# Volunteering at a Student-Run Clinic and Matching into Primary Care Specialties

**DOI:** 10.64898/2026.01.22.26344668

**Authors:** Daniel C. Brock, Akshat Kumar, Hannah Engebretson, Sarah Grant, Zuha Khan, Panayiotis D. Kontoyiannis, Michael J. DiLeo, Spoorthi Kamepalli, Mackenzie K. Joe, Nicholas Peoples, Michael A. Altman, Malford T. Pillow, Dana L. Clark

**Author notes:** Correspondence: D.L.C.

## Abstract

**Background:** Student-run clinics (SRC) serve a unique role in healthcare by addressing the needs of underserved communities while providing medical students hands-on learning experiences. The Houston Outreach Medicine Education and Social Services (HOMES) Clinic is a SRC and a program of Healthcare for the Homeless - Houston that provides medical care to individuals experiencing unstable housing in Houston, Texas. Amid a growing shortage of primary care physicians in the United States, understanding factors that influence specialty choice is critical. This study aimed to explore whether volunteering at HOMES Clinic is associated with an increased likelihood of matching into primary care specialties.

**Methods:** This study used a retrospective cohort design of HOMES Clinic volunteers from 2014-2025. Students who volunteered at HOMES Clinic represented the exposure group (n=1,157), while non-volunteers served as the reference group (n=3,666). The primary outcome was the association between volunteering and matching into primary care specialties. Secondary outcomes included residency program rank, in-state residency placement, and induction into the Alpha Omega Alpha and Gold Humanism Honor Societies.

**Results:** HOMES Clinic volunteers matched into primary care specialties at a 7.5% higher rate than non-volunteers (p=1.3×10^-5^; OR=1.38; 95% CI = 1.20-1.59). Conversely, HOMES volunteers showed a 5.3% lower proportion of students who matched into surgical specialties (p=8.9×10^-4^; OR=0.76; 95% CI = 0.65-0.89). Volunteers also showed a modest association with matching into higher-ranked residency programs (p<0.05) and had 25% higher odds of Alpha Omega Alpha induction and 41% higher odds of Gold Humanism Honor Society induction.

**Conclusions:** Volunteering at HOMES Clinic showed a positive association with matching into primary care specialties. This trend likely reflects both self-selection of students interested in primary care and the influence of SRC experience on shaping student residency specialty choices. Our results provide insights into how medical schools and SRCs foster the development of the next generation of primary care physicians.

## Introduction

Student-run clinics (SRCs) are an integral part of medical education, with over 208 operating within institutions of the Association of American Medical Colleges (AAMC) and at least 75% of US medical schools having one or more SRC^1^. SRCs serve a dual role by addressing the health needs of underserved communities while offering medical students hands-on learning opportunities. It has been hypothesized that volunteering at SRCs, particularly those emphasizing holistic and preventive care, may influence medical students toward careers in primary care^2–6^. This is especially important given the ongoing shortage of primary care physicians in the United States, which is projected to grow substantially over the upcoming decades^7,8^.

The National Resident Matching Program (NRMP) pairs graduating medical students with residency programs to train future specialists^9^. The match process is highly competitive and many factors can affect an applicant’s success, including academic performance, letters of recommendation, research, nominations to honor societies, and community service^10,11^. Participation in SRCs offers students a unique opportunity to distinguish themselves by demonstrating a commitment to honing clinical skills, engaging with diverse patient populations, and developing a deeper understanding of the social determinants of health. Additionally, volunteering is an important criterion in the selection process for honor societies, such as Alpha Omega Alpha (AOA) and Gold Humanism Honor Society (GHHS), both of which can significantly influence residency applications^12–15^. Given the rich learning environment provided by SRCs, it is important to examine the relationship between SRC volunteering and the residency match to understand how experiential learning may shape students’ career trajectories.

Previous research on SRCs and residency match outcomes have yielded variable associations regarding SRC volunteering and specialty choice. In two studies of SRC volunteers, there was no significant association between SRC volunteering and pursuit of primary care specialties^2,16^. Similarly, a literature review of ten studies reported no significant correlation between SRC volunteering and selection of specific residency specialties^17^. In contrast, three other studies found positive correlations between volunteering at a SRC and future primary care practice^18–20^. An additional study found significant associations between student participation in specialty-specific community health clinics and matching into related specialties, such as dermatology and neurology, as well as a positive association between primary care clinic volunteering and matching into primary care residencies^3^. These conflicting findings highlight the need for further research to clarify the relationship between SRC involvement and residency outcomes.

The Houston Outreach Medicine Education and Social Services (HOMES) Clinic^21^ is a SRC and a program of Healthcare for the Homeless – Houston (HHH)^22^ that provides medical care to individuals experiencing unstable housing in Houston, Texas. Unlike many SRCs that draw exclusively from a single institution, HOMES accepts student volunteers from three medical schools: Baylor College of Medicine, McGovern Medical School at UTHealth Houston, and the Tilman J. Fertitta Family College of Medicine at the University of Houston. This multi-institutional structure strengthens the generalization of the findings from HOMES Clinic, supporting associations that reflect the experiences of students rather than institution-specific factors. As such, HOMES Clinic is uniquely positioned to contribute to the evolving conversation on SRC participation and medical student specialty choice. Here, we explored the relationship between volunteering at HOMES Clinic and residency match outcomes, nominations to honor societies, and the likelihood of pursuing primary care specialties. This study represents the largest individual analysis of SRC volunteer outcomes to date, examining both specialty choice and match characteristics. Insights from this work may guide mentorship, enhance training opportunities for SRC volunteers, and support broader institutional efforts to integrate SRCs into medical education.

## Methods

### Study Setting

HOMES Clinic is a multi-institutional SRC, a program of Healthcare for the Homeless Houston (HHH)^22^, that provides medical care to unhoused individuals in downtown Houston, Texas. HOMES Clinic welcomes student volunteers from Baylor College of Medicine (BCM), McGovern Medical School at UTHealth Houston (UTH), University of Houston Tilman J. Fertitta Family College of Medicine, and the University of Houston College of Pharmacy. Patients are eligible to receive care based on documented homeless status, aligning with guidelines established by the Health Resources and Services Administration and the Bureau of Primary Care.

Medical students volunteer in a variety of roles depending on their progress in their respective fields: manager, manager-in-training, clinical student, pre-clinical student, pharmacy student, or health advocate. Managers are students who ensure that clinic procedures are in line with standard operating procedures. Manager-in-training volunteers assist managers and receive training to become managers. Clinical students are medical students who have entered their clinical clerkships and are responsible for patient documentation and leading the clinical encounter. Pre-clinical students are medical students in their basic science training phase who conduct the history and physical exam under the supervision of the clinical student. Pharmacy students are in their second to fourth year of pharmacy graduate training and gather medication history, provide medication counseling to patients, and track pharmacy inventory. Health advocate volunteers are students who function as social workers by connecting clients to resources using a structured social resource guide developed specifically for HOMES Clinic^23^. All clinical care is ultimately closely supervised by a faculty attending physician.

Volunteer teams at HOMES Clinic consist of two pre-clinical students, one pharmacy student, and one clinical student (**Figure 1**). The day begins in The Beacon^24^, a space where individuals experiencing homelessness can access services including meals, laundry, and showers. Pre-clinical and pharmacy volunteers identify individuals with healthcare needs at their tables in The Beacon. Patients with social concerns are referred to health advocate volunteers. Patients with medical needs are directed to clinical students who triage patients and assess whether their needs can be addressed in the clinic. After triage, volunteer teams move to the clinic where they conduct holistic patient visits, including vital signs, history and physical exams, and patient discharge. Students collaborate to create differential diagnoses and work with attending physicians to formally diagnose and treat patients.

**Figure 1:**
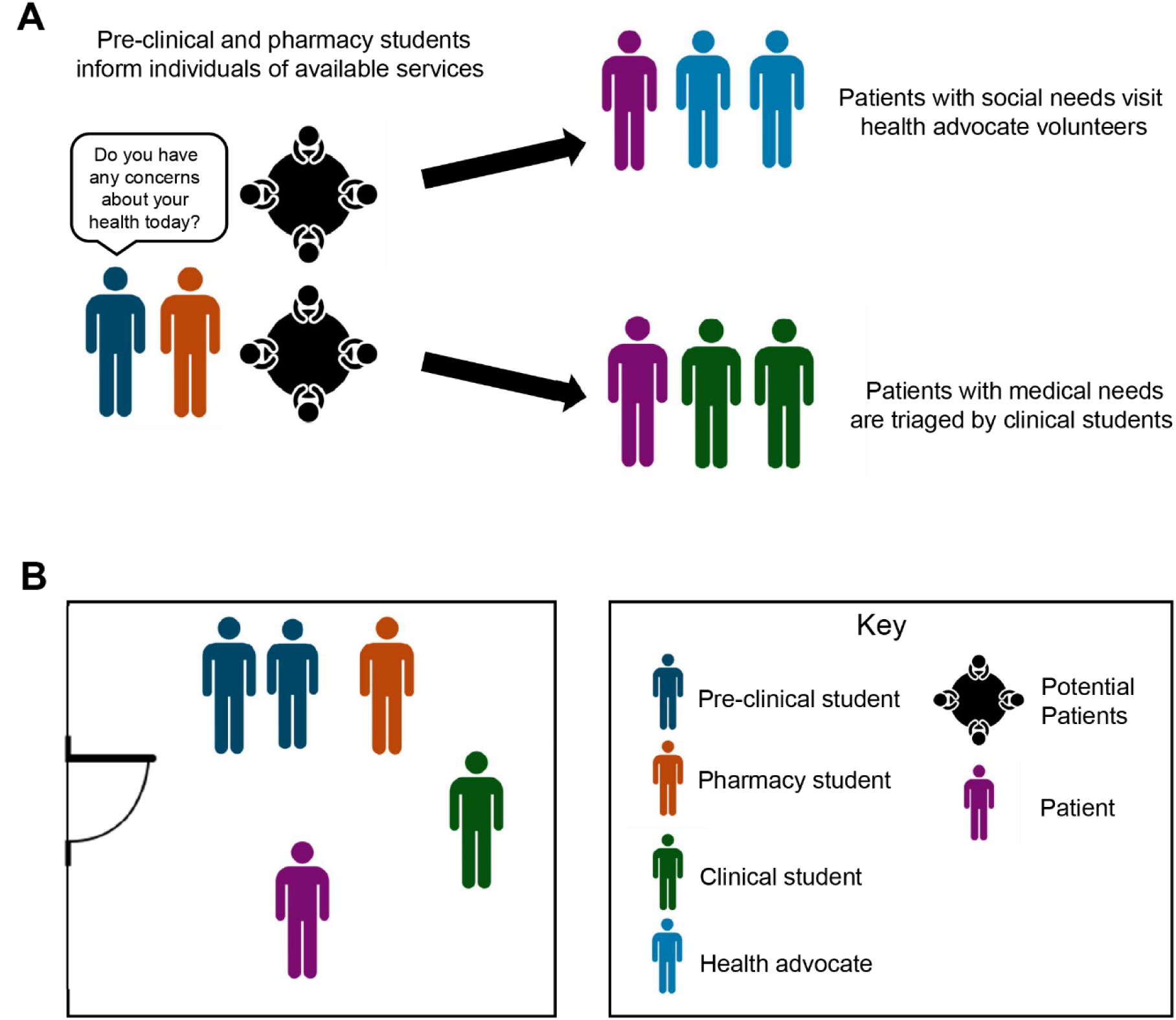
Student Flow during HOMES Clinic. A) Assessment of clients at their tables in the Beacon, including visits with health advocates for social needs and clinical students for medical needs. B) Clinical team consisting of two pre-clinical students, one pharmacy student, and one clinical student per patient.

### Study Design

We conducted a retrospective cohort study to evaluate medical student volunteers at HOMES Clinic between 2014 and 2025. Eligible HOMES Clinic volunteers included medical students from BCM and UTH. The Tilman J. Fertitta Family College of Medicine at the University of Houston (UH) enrolled its inaugural class in 2020, and volunteering at HOMES began in 2022; thus, we did not include UH in our analysis because no graduating students were available during the study period. We compiled annual HOMES Clinic attendance registries and obtained residency match lists from the deans of student affairs at BCM and UTH. We designated students who volunteered at HOMES Clinic as the exposure group and labeled non-volunteers as the reference group.

### Data Collection and Eligibility

This study was reviewed by the Institutional Review Boards (IRB) at BCM and UTH. The IRB at BCM approved of this study (IRB number: H-56505) and the IRB at UTH determined it to be exempt. Both BCM and UTH approved of data collection and use of match results.

Medical students at BCM and UTH received monthly emails inviting them to sign up for volunteer positions at HOMES Clinic. The HOMES Clinic executive board recorded signup information in annual volunteer attendance registries. The registries contained student names, volunteering date, medical school affiliation, volunteer roles, and absence status. We manually reviewed signup sheets for typos, filtered for absent volunteers, and merged attendance registries into a single dataset spanning 2014-2025.

We also compiled annual residency match lists from BCM and UTH into a single dataset. These lists included student names, graduation years, matched specialties, residency programs, and honor society nominations to Alpha Omega Alpha (AOA) and Gold Humanism Honor Society (GHHS). To pair volunteers with their corresponding residency match data, we used student names and school affiliations from the HOMES Clinic volunteer registry as unique identifiers. Fuzzy string matching was performed using the “thefuzz” python package to account for potential discrepancies in name formatting. A custom script automatically paired exact name matches between the two datasets and prompted user input for pairing inexact names, ranked by similarity metrics.

Students classified as volunteers if they participated in at least one HOMES Clinic volunteer session. We placed students who never volunteered in the non-volunteer reference group. All students included in the analysis matched into an Accreditation Council for Graduate Medical Education (ACGME)-accredited U.S. residency program. We excluded students who did not match (n=78), and those who deferred or did not apply to residency. Additionally, we excluded students from downstream analyses if they matched into preliminary or transitional programs unless we could identify a definitive residency specialty.

To determine the relationship between HOMES Clinic volunteering and residency program rank, we incorporated specialty-specific residency rankings from the Doximity Residency Navigator^25^ (accessed on December 18, 2024). Doximity ranked residency programs based on reputation, research output, and program size.

### Outcomes

We measured the proportion of HOMES Clinic volunteers who matched into primary care specialties as the main outcome of this study. Primary care constituted the sum of internal medicine, pediatrics, family medicine, and internal medicine-pediatrics. Similar to a prior report from the AAMC^26^, we categorized the remaining specialties into the following groups: surgical (general surgery, neurological surgery, obstetrics & gynecology, ophthalmology, orthopedic surgery, otolaryngology, plastic surgery, thoracic surgery, urology, and vascular surgery), procedural (anesthesiology, dermatology, emergency medicine, radiation oncology, and interventional radiology), and other (neurology, child neurology, psychiatry, diagnostic radiology, pathology, medical genetics, and physical medicine & rehabilitation). Next, we further compared the proportion of HOMES volunteers versus non-volunteers for individual specialties, restricted to specialties with a minimum of five matched students per group. Lastly, we evaluated other volunteer factors that may influence specialty choice, including the total number of volunteer shifts, length of time volunteered, volunteer role, and affiliated medical school.

Secondary outcomes included residency program rankings, in-state residency, and induction into honor societies among HOMES Clinic volunteers versus non-volunteers. We defined in-state residency as attending a residency program located in Texas. Lastly, we used induction lists for AOA and GHHS to examine if HOMES Clinic participation was associated with higher rates of election to academic or humanism-based honor societies.

### Statistical Analysis

We utilized Fisher’s exact test to compare the proportion of students who matched into primary care specialties between HOMES Clinic volunteers and non-volunteers. For specialty-specific analyses, Fisher’s exact test compared HOMES volunteers versus non-volunteers for each specialty, followed by correction for multiple comparisons using the false discovery rate (FDR). We report adjusted p-values and odds ratios with 95% confidence intervals. We also used Fisher’s exact test to compare the proportions of students matching into in-state residency programs and the proportions inducted into AOA and GHHS.

For continuous variables, we assessed normality using the Shapiro-Wilk test and the DHARMa package in R. We determined that our data was not normally distributed, so we used non-parametric tests, such as Wilcoxon signed rank test to evaluate differences in residency program rank and size. We conducted multiple logistic regression to determine the influence of volunteer metrics on the proportions of students choosing primary care, such as volunteer frequency, length of time volunteered, volunteer role, and affiliated medical school.

We performed data analysis using Python (v.3.12.3) and R (v.4.2.2). Python packages utilized include pandas (v.2.2.2), numpy (v.1.26.4), and thefuzz (v.0.22.1). R packages include the tidyverse suite of packages (v.2.0.0), plyr (v.1.8.9), plotrix (v.3.8-4), finalfit (v.1.0.8), rstatix (v.0.7.2), performance (v.0.13.0), DHARMa (v.0.4.7), ggeffects (v.2.2.1), ggforestplot (v.0.1.0), ggpubr (v.0.6.0) and patchwork (v.1.3.0). The code used to generate figures and statistics can be found on our publicly available GitHub repository: https://github.com/HOMES-Clinic/volunteer_outcomes_study.git

## Results

### Study Population

We identified 1,667 unique medical student volunteers in the HOMES Clinic volunteer registry (**Figure 2**). The residency match dataset included 4,901 medical students, of whom 4,823 successfully matched into residency. Of the HOMES Clinic volunteers, 1,157 (69.4%) matched into residency; most unmatched volunteers were current students who had not yet applied to residency. We could not confidently link 30 (1.8%) students, likely due to potential name changes during medical school or discrepancies in the volunteer sign-up records. Among matched students, 3,666 did not volunteer at HOMES Clinic, who served as the reference group. The volunteer group consisted of 577 students from BCM (49.9%) and 580 from UTH (50.1%) (**Table 1**). Pre-clinical volunteers represented the most common volunteer position (46.9%), followed by clinical students (30%), managers-in-training (11.1%), managers (9.4%), and health advocates (2.6%) (**Table 1, Supplemental Figure 1)**. Managers exhibited the highest volunteer frequency (mean = 3.64, SEM = 0.29), while health advocate volunteers showed the lowest (mean = 1.48, SEM = 0.11).

**Figure 2:**
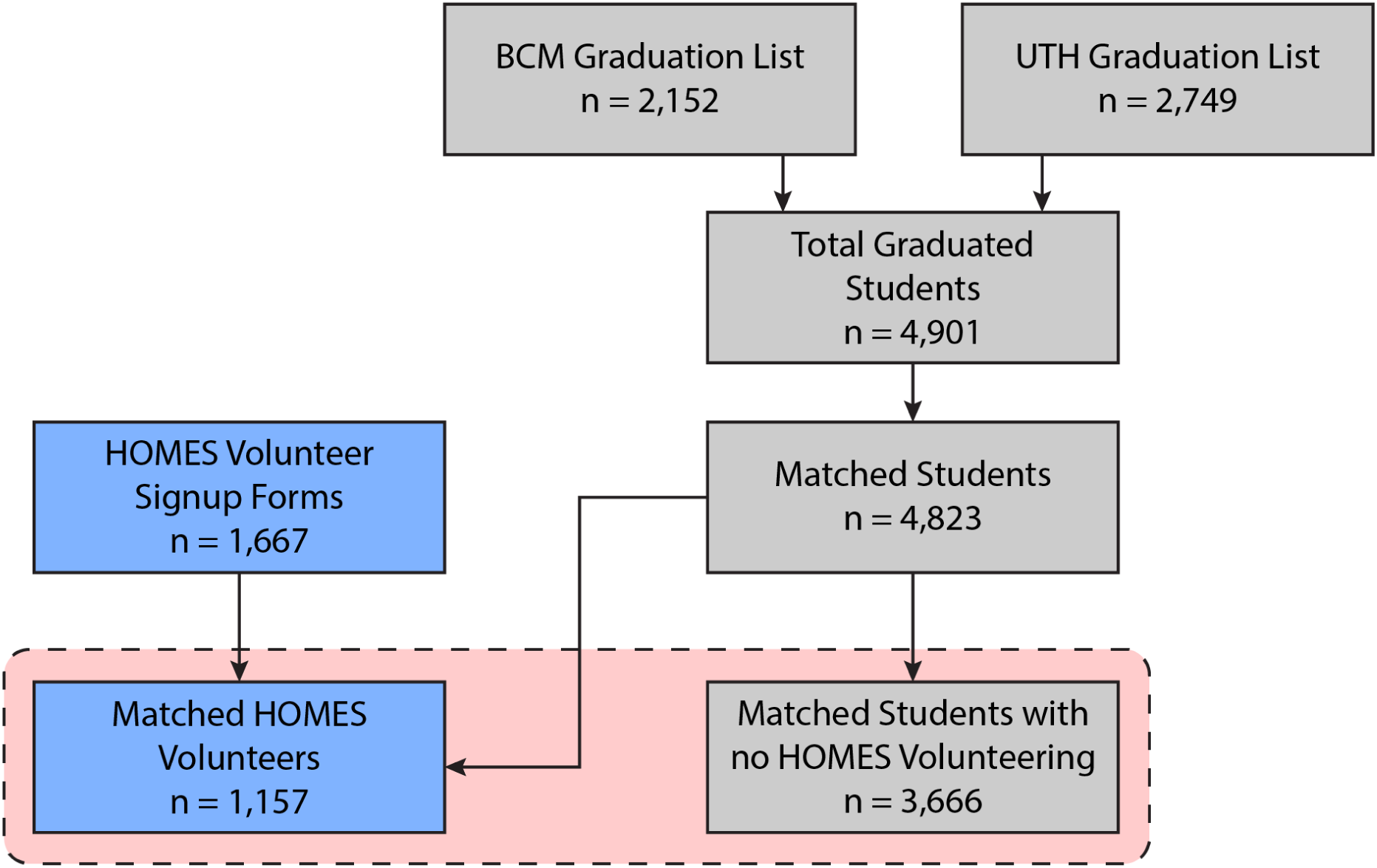
Study Participants. Data sources are color-coded with blue indicating HOMES Clinic volunteer attendance records and grey denoting medical school residency match lists. The red dashed box highlights the primary comparison of interest: matched students who volunteered at HOMES Clinic compared to matched students who did not volunteer.

**Table 1:**
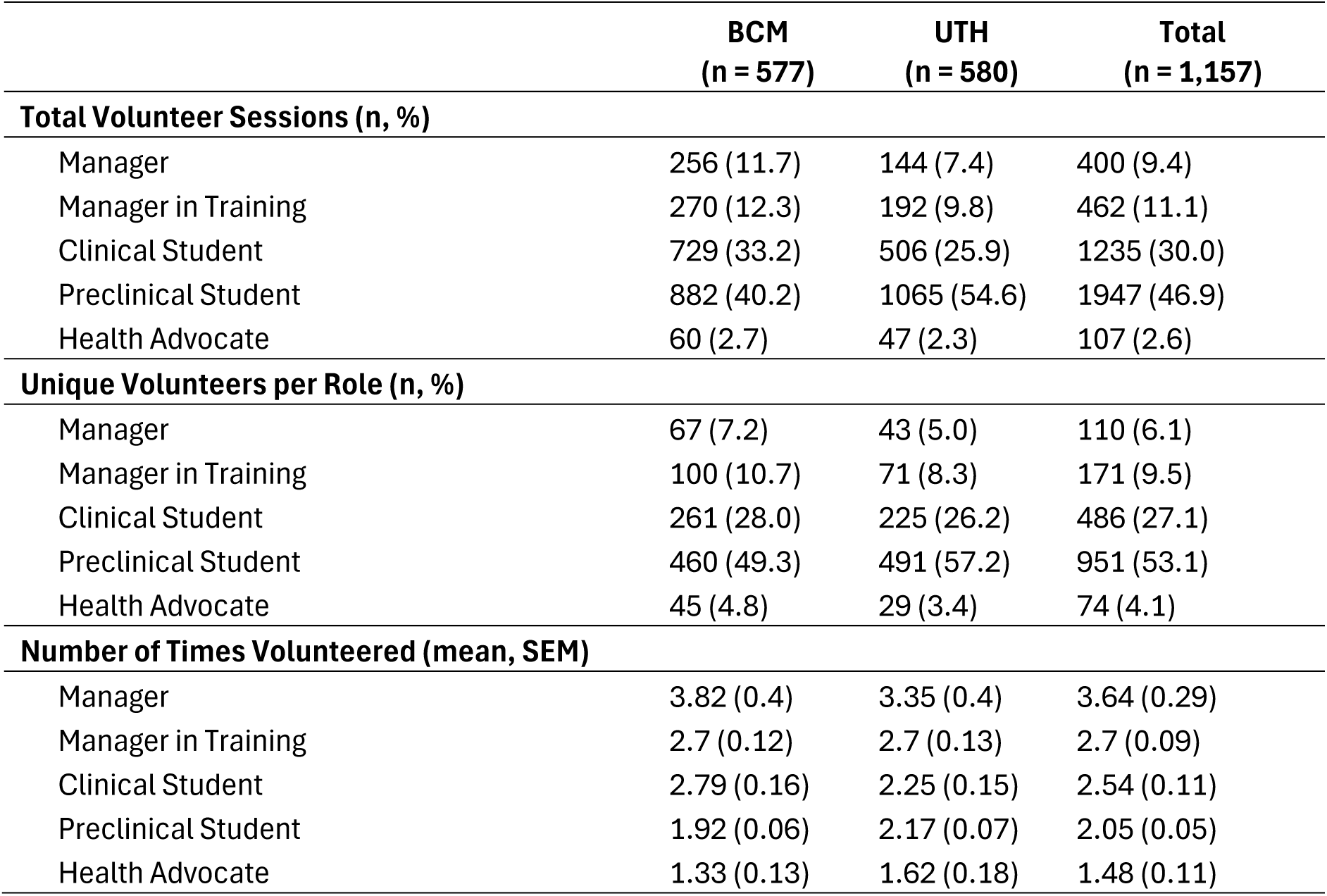
HOMES Clinic Volunteers. Student Volunteers from 2014-2025

|  | BCM<br>(n = 577) | UTH<br>(n = 580) | Total<br>(n = 1,157) |
| --- | --- | --- | --- |
| **Total Volunteer Sessions (n, %)** |  |  |  |
| Manager | 256 (11.7) | 144 (7.4) | 400 (9.4) |
| Manager in Training | 270 (12.3) | 192 (9.8) | 462 (11.1) |
| Clinical Student | 729 (33.2) | 506 (25.9) | 1235 (30.0) |
| Preclinical Student | 882 (40.2) | 1065 (54.6) | 1947 (46.9) |
| Health Advocate | 60 (2.7) | 47 (2.3) | 107 (2.6) |
| **Unique Volunteers per Role (n, %)** |  |  |  |
| Manager | 67 (7.2) | 43 (5.0) | 110 (6.1) |
| Manager in Training | 100 (10.7) | 71 (8.3) | 171 (9.5) |
| Clinical Student | 261 (28.0) | 225 (26.2) | 486 (27.1) |
| Preclinical Student | 460 (49.3) | 491 (57.2) | 951 (53.1) |
| Health Advocate | 45 (4.8) | 29 (3.4) | 74 (4.1) |
| **Number of Times Volunteered (mean, SEM)** |  |  |  |
| Manager | 3.82 (0.4) | 3.35 (0.4) | 3.64 (0.29) |
| Manager in Training | 2.7 (0.12) | 2.7 (0.13) | 2.7 (0.09) |
| Clinical Student | 2.79 (0.16) | 2.25 (0.15) | 2.54 (0.11) |
| Preclinical Student | 1.92 (0.06) | 2.17 (0.07) | 2.05 (0.05) |
| Health Advocate | 1.33 (0.13) | 1.62 (0.18) | 1.48 (0.11) |

### Residency Match Outcomes for HOMES Volunteers

Among 1,157 HOMES volunteers, 477 (41.2%) matched into primary care specialties, compared to 1,234 out of 3,666 non-volunteers (33.7%), representing a 7.5% higher proportion of HOMES volunteers matching into primary care (p=1.3×10^-5^; Odds Ratio (OR) = 1.38; 95% Confidence Interval (CI) = 1.20-1.59) (**Figure 3A, Supplemental Table 1**). Conversely, HOMES volunteers displayed a 5.3% lower proportion of students who matched into surgical specialties (p=8.9×10^-4^; OR=0.76; 95% CI = 0.65-0.89). HOMES volunteers did not differ relative to non-volunteers with respect to procedural or “other” specialties (definitions in Methods).

**Figure 3:**
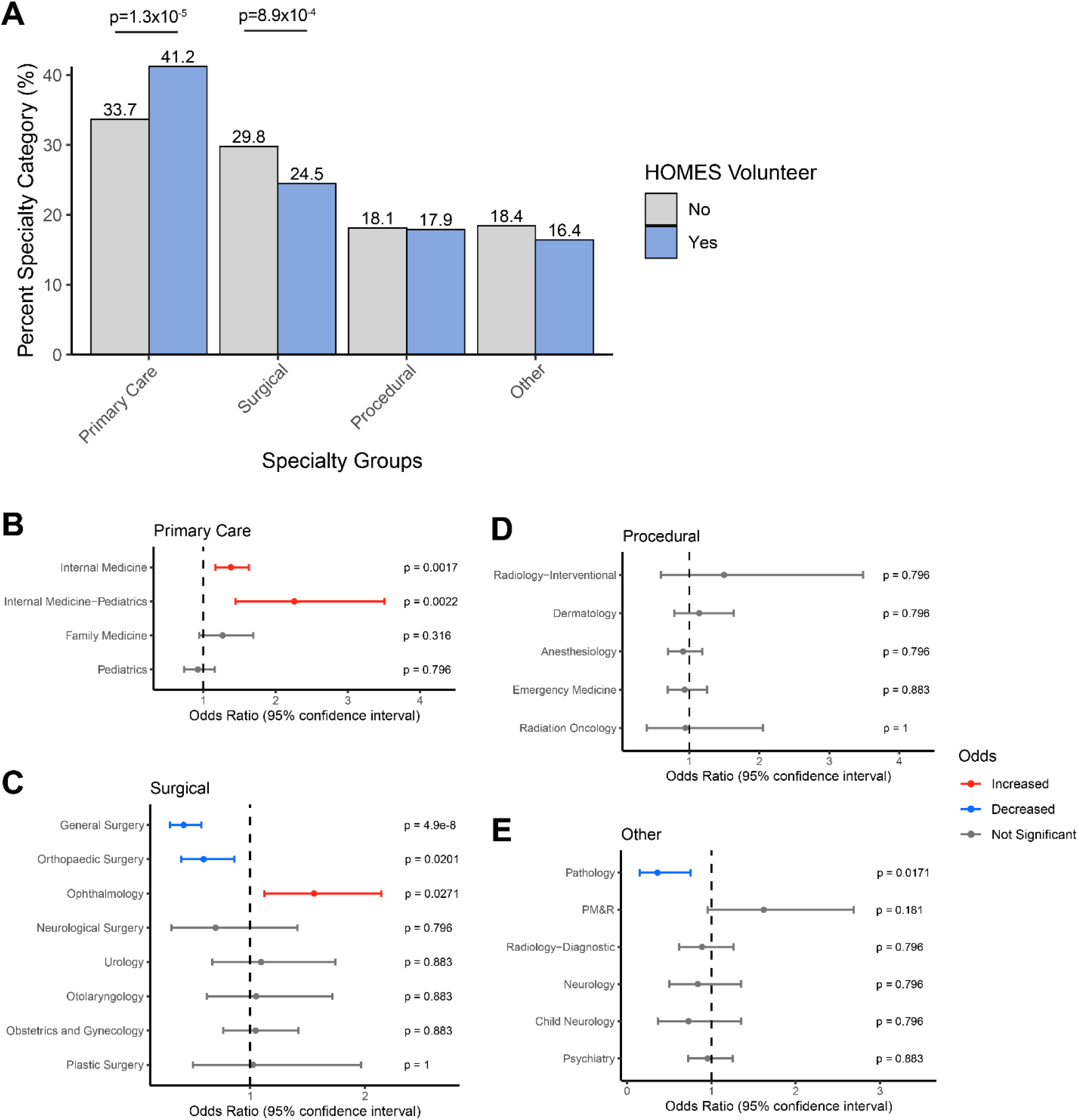
Match Results of HOMES Volunteers Compared with Non-Volunteers. A) Proportions of specialty groups for HOMES volunteers compared to non-volunteers. B) Specialty-specific odds for primary care, C) surgical, D) procedural, and E) other specialties. The central point represents the odds ratio and the lower and upper error bars represent the 95% confidence interval. Positive associations are colored red and negative associations are colored blue. Adjusted p-values are shown to the right side of each graph.

To assess which individual specialties drove these changes, we disaggregated specialty groups into individual specialties and tested for over- or underrepresentation of HOMES volunteers. HOMES volunteers showed a higher proportion of students who matched into internal medicine (p=0.0017; OR=1.38; 95% = 1.18-1.63) and internal medicine-pediatrics (p=0.0022; OR=2.26; 95% CI = 1.44-3.50) (**Figure 3B; Supplemental Table 2**). For surgical specialties, HOMES volunteers showed a lower proportion of students who matched into general surgery (p=4.9×10^-8^; OR=0.42; 95% CI = 0.30-0.57) and orthopedic surgery (p=0.02; OR=0.59; 95% CI = 0.40-0.86) (**Figure 3C**). In contrast, HOMES volunteers displayed a higher proportion of students matching into ophthalmology (p=0.027; OR=1.56; 95% CI = 1.12-2.14). HOMES volunteers did not differ in proportions for procedural specialties (**Figure 3D**). Lastly, HOMES volunteers showed a lower proportion of students who matched into pathology (p=0.017; OR=0.36; 95% CI = 0.15-0.75) (**Figure 3E**).

To determine if medical students’ affiliate medical schools influenced specialty outcomes, we performed the same analysis, split by students’ affiliate medical school. Students from UTH drove the positive association between HOMES volunteers and internal medicine (p=0.013; OR=1.46; 95% CI = 1.15-1.84), while students from BCM drove the association with internal medicine-pediatrics (p=0.01; OR=2.85; 95% CI = 1.52-5.39) (**Supplemental Figure 2; Supplemental Table 3**). UTH students drove the negative association for general surgery (p=6.04×10^-8^; OR=0.3; 95% CI = 0.18-0.47) and the positive association with ophthalmology (p=0.014; OR=2.12; 95% CI = 1.29-3.45). Specialties such as orthopedic surgery and pathology became significant only after combining the effects of students from both BCM and UTH.

### Residency Program Rank for HOMES Volunteers

Next, we utilized specialty-specific residency ranks gathered from the Doximity Residency Navigator^25^ to determine if HOMES volunteers matched into more competitive residency programs. HOMES volunteers showed a modest association of matching into higher ranked programs, ranked by reputation (p=0.017), and a larger association of matching into higher ranked programs ranked by research output (p=0.002; **Table 2**). HOMES volunteers did not show a significant association of matching into larger residency programs (p=0.072). Geographically, HOMES volunteers did not differ from non-volunteers for matching into in-state residency programs (p=0.15; **Supplemental Table 4**). HOMES Volunteers also did not show any associations for matching into specific ERAS regions compared to non-volunteers.

**Table 2:** Residency program ranks. Lower numbers indicate higher ranked residency programs

|  |  | n, Control<br>(n = 3666) | n, HOMES<br>(n = 1157) | p-value |
| --- | --- | --- | --- | --- |
| Rank: Reputation | Median (IQR) | 37.0 (17.0 to 69.5) | 35.0 (16.0 to 64.0) | 0.017 |
| Rank: Research | Median (IQR) | 43.0 (20.0 to 82.0) | 37.0 (19.0 to 64.0) | 0.002 |
| Program Size | Median (IQR) | 48.0 (25.0 to 84.0) | 51.0 (25.0 to 107.8) | 0.072 |

### Honor Society Membership Among HOMES Clinic Volunteers

Volunteering at HOMES Clinic can serve as an impactful community service experience and leadership experience for those who pursue manager training. Thus, we analyzed if volunteering at HOMES Clinic was associated with different likelihoods of induction into honor societies at the time of graduation. 20.1% of HOMES volunteers showed induction to the Alpha Omega Alpha (AOA), compared to 16.8% for non-volunteers (p=0.011; OR=1.25; 95% CI = 1.05-1.48) (**Figure 4A**; **Supplemental Table 5**). HOMES volunteers had a greater likelihood of being inducted into the Gold Humanism Honor Society (GHHS), with a 16.3% chance of induction, compared to 12.1% for non-volunteers (p=9.3×10^-4^; OR=1.41; 95% CI = 1.15-1.73) (**Figure 4B**).

**Figure 4:**
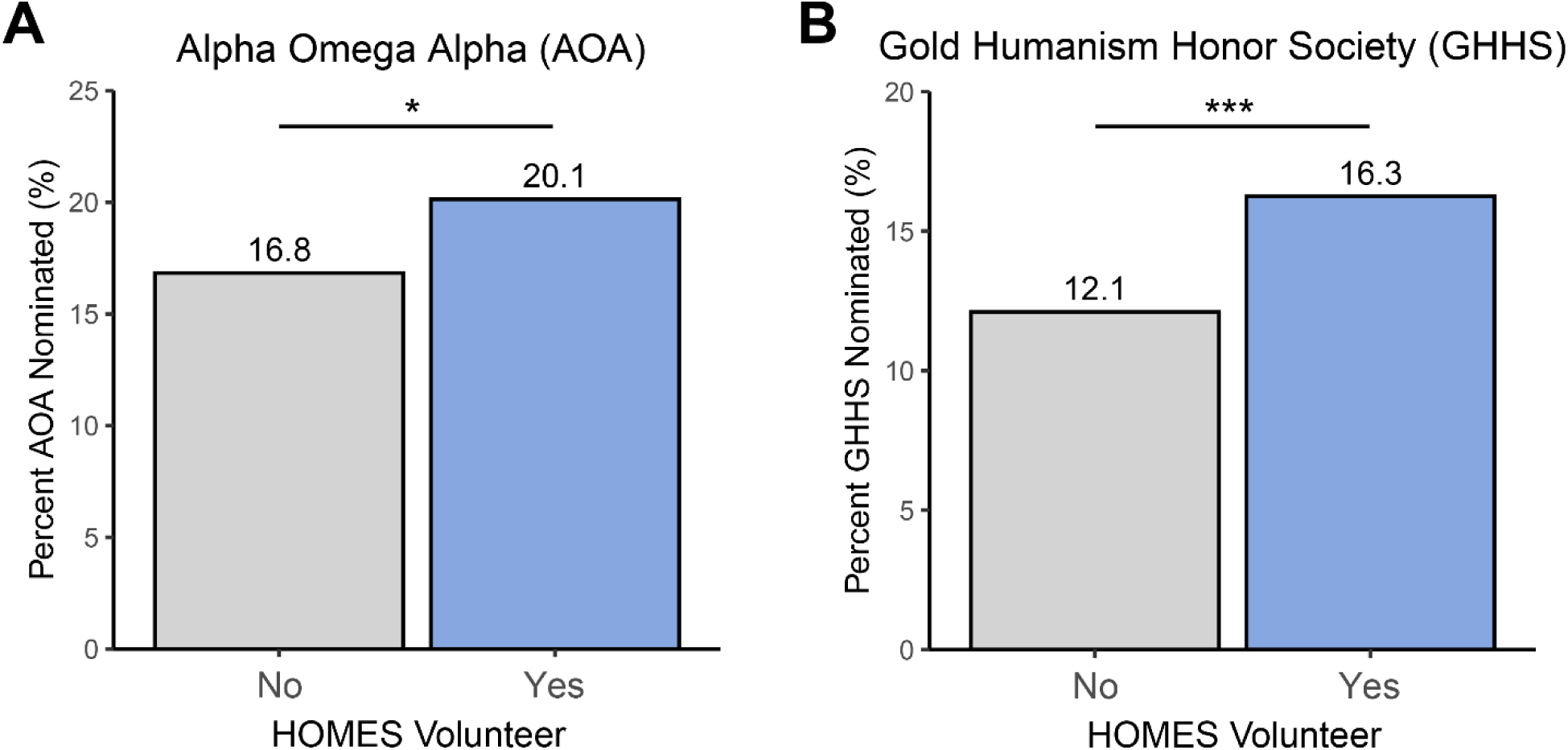
HOMES Volunteers Show Higher Likelihood of Induction into Honor Societies. A) Proportions of students nominated to Alpha Omega Alpha (AOA) honor society and B) Gold Humanism Honor Society (GHHS). * p < 0.05, *** p < 0.001.

### Influence of Volunteer Frequency and Role on Pursuing Primary Care Specialties

We next performed multivariate logistic regression to examine how effector variables influenced the proportion of HOMES Clinic volunteers pursuing primary care specialties. Effector variables included volunteer frequency, length of time volunteered, and volunteer role. Volunteer frequency, length of time volunteering, volunteer role, and affiliated medical school did not significantly predict if HOMES volunteers pursued primary care specialties (**Table 3**).

**Table 3.** Multivariate Logistic Regression Assessing Volunteer Factors for Matching into Primary Care Specialties. The reference level for volunteer role was preclinical volunteers and the reference for affiliate medical school was BCM.

|  | Odds Ratio [95% CI] | p value |
| --- | --- | --- |
| Volunteer Frequency | 0.97 [0.92, 1.03] | 0.35 |
| Length of Time Volunteered | 1.01 [0.99, 1.03] | 0.23 |
| Role: Health Advocate | 0.74 [0.37, 1.43] | 0.39 |
| Role: Clinical Student | 1.06 [0.79, 1.42] | 0.70 |
| Role: Manager in Training | 0.86 [0.49, 1.5] | 0.60 |
| Role: Manager | 1.58 [0.84, 3] | 0.16 |
| School: UTH | 0.88 [0.7, 1.12] | 0.30 |

In contrast, we found that volunteer frequency positively associated with volunteers who matched into surgical specialties (p=0.011; OR=1.08; 95% CI = 1.02-1.15) while role as a manager was negatively correlated with volunteers who matched into surgical specialties (p=0.015; OR=0.37; 95% CI = 0.16-0.80) (**Supplemental Table 6**). Among HOMES Clinic volunteers, medical school affiliation with UTH was associated with matching into residency programs 16.5 positions lower than volunteers from BCM (p=7.33×10^-4^; β = 16.5; 95% CI = 7.87 to 25.12) (**Table 4**). Additional analyses separating residency program reputation and research output rankings are presented in **Supplemental Table 7**.

**Table 4:** Multivariate Spearman Correlation Evaluating Volunteer Factors Associated with Residency Program Competitiveness. Negative β Coefficients indicate higher ranked programs (e.g. rank #1 compared to rank #27). The reference level for volunteer role was preclinical volunteers and the reference for affiliate medical school was BCM.

| | $\beta$ Coefficient [95% CI] | p value |
| --- | --- | --- |
| Volunteer Frequency | -0.01 [-0.05, 0.03] | 0.90 |
| Length of Time Volunteered | 0.02 [-0.02, 0.06] | 0.70 |
| Role: Health Advocate | 0.51 [-24.26, 25.27] | 0.97 |
| Role: Clinical Student | -4.39 [-15.06, 6.28] | 0.84 |
| Role: Manager in Training | -1.97 [-23.29, 19.35] | 0.97 |
| Role: Manager | -2.44 [-21.93, 17.04] | 0.97 |
| School: UTH | 16.5 [7.87, 25.12] | 7.33E-04 |

## Discussion

Conflicting evidence exists regarding whether SRC participation during medical school influences specialty choice^5,6,17,19,20^. To address this gap, we conducted the largest single-cohort study to date, spanning 11 years across two medical schools. We found that SRC volunteers were significantly more likely to enter primary care specialties compared with non-volunteers. Volunteering was also associated with matching into higher-ranked residency programs and increased rates of induction into honor societies such as AOA and GHHS. A recent meta-analysis by Peoples et al.^27^, which combined the results of this study with six other studies, found that volunteering at SRCs was associated with a higher likelihood of pursuing a primary care specialty. These findings provide new insights into how SRC participation may shape residency outcomes and highlight the potential role of volunteer experiences in supporting students’ career development.

The primary outcome of this study demonstrated that a significantly higher proportion of HOMES Clinic volunteers matched into primary care specialties. Within primary care, internal medicine and internal medicine-pediatrics emerged as the main drivers of this association. This pattern likely reflects two factors: (1) self-selection of students already inclined toward primary care and (2) the influence of SRC volunteering on students’ specialty choices. There exists a large overlap in scope and function between the HOMES Clinic and primary care, as SRCs typically emphasize primary and preventive care^28^. While these results support the hypothesis that SRC participation can shape specialty decisions, prospective studies are needed to establish a causal relationship.

Most studies examining SRC volunteer outcomes have focused on primary care specialties, with limited evaluation of other medical fields^29^. Our study also analyzed the relationship between SRC participation and matching into non-primary care specialties, and we found that HOMES volunteers showed an overall trend of reduced likelihood of choosing surgical specialties. These findings underscore the importance of expanding SRC research to include specialties beyond primary care, thereby broadening understanding of how SRC participation influences the full spectrum of medical specialty choices.

An exception to the overall trend in surgical specialties is the positive association between HOMES volunteers and matching into ophthalmology. This outcome likely reflects HOMES Clinic’s development of specific ophthalmology service opportunities in response to the disproportionate burden of vision-related conditions among homeless populations^30,31^. HOMES Clinic established a weekly ophthalmology clinic, focused on screening, prevention, and referral, providing students with structured exposure, mentorship, and vocational identity development in this field. Given that mentorship is a well-documented factor influencing surgical career choice^32–35^, the absence of comparable non-ophthalmologic surgical opportunities may contribute to students interested in other surgical fields seeking alternative avenues for mentorship.

Our findings underscore the importance of specialty representation at SRCs and the potential of expanding SRC volunteer opportunities to encompass a broader range of specialties. However, recommendations to expand specialty services at SRCs must acknowledge the complexities and challenges involved^36^. Specialty clinics raise important ethical and logistical considerations, including ensuring adequate multi-disciplinary care, establishing competency standards for students, and addressing liability protections^37^. Thus, while expansion of specialty representation offers educational opportunities, future efforts should be guided by careful consideration of these guidelines.

Examining honor society membership demonstrated that HOMES Clinic volunteers were more likely to be inducted into AOA and GHHS compared with non-volunteers. Both AOA and GHHS emphasize service, leadership, and compassionate patient care, suggesting that volunteering at HOMES Clinic provides meaningful opportunities to develop and demonstrate these qualities. These results are consistent with prior studies showing a positive association between SRC participation and honor society induction^15,38^. Exposure to HOMES Clinic’s patient population, composed of individuals experiencing homelessness, heightens awareness of the social determinants of health. This visibility promotes discussion of complex social factors and fosters a deeper appreciation for their impact on patient care^28^. Furthermore, manager training and the leadership responsibilities of clinical students at HOMES Clinic are additional opportunities to develop skills valued by both AOA and GHHS.

This study has several limitations. Its retrospective design introduces inherent selection bias, and causal relationships cannot be determined. Although SRC participation may influence students to pursue primary care, it is equally plausible that students with preexisting interest in primary care are more likely to volunteer. Because specialty choice is multifactorial^39^, future work will track volunteers’ specialty preferences longitudinally to assess factors that influence changes in preference over time. In the absence of a standardized, objective residency ranking system, we used Doximity rankings as a surrogate for program reputation and research output. These rankings are derived from surveys of board-certified physicians on the Doximity platform but omit objective measures such as board pass rates, fellowship match rates, and research funding^25,40^. Despite these methodological limitations, Doximity rankings demonstrably influence applicant decision-making and program reputation^40,41^. In one survey of 2,152 applicants, 62% reported using Doximity during their application cycle, and 79% of these indicated that the rankings affected their rank lists^41^. Thus, although Doximity’s methodology has significant limitations, its clear influence on residency decision-making supports its use as a relevant ranking metric in this study.

In conclusion, our multi-institution retrospective analysis of 4,823 medical students found that a higher proportion of SRC volunteers matched into primary care specialties compared to non-volunteers. Sub-analyses further revealed that SRC volunteers had higher rates of induction into medical honor societies and were more likely to match into higher-ranked residency programs. These findings suggest that volunteering at an SRC may help shape medical students’ professional identity and provide a valuable opportunity to explore clinical interests, potentially influencing their career trajectories.

## Data Availability

All data produced in the present study are available upon reasonable request to the authors.

https://github.com/HOMES-Clinic/volunteer_outcomes_study

## Ethics Statement

This study was reviewed by the Institutional Review Boards (IRB) at Baylor College of Medicine and McGovern Medical School. IRB at Baylor College of Medicine approved this study (IRB number: H-56505) and the IRB at McGovern Medical School determined this study to be exempt.

## Acknowledgements

We thank Kathryn Rogers at the Healthcare for the Homeless Houston (HHH) for supporting this study and providing feedback while drafting this manuscript. We also thank Dr. Hugh Stoddard for providing feedback on data analysis and manuscript writing.

## Author Contributions

Authors contributed to this work in the following capacities: Conceptualization: D.C.B., A.K., S.K.; methodology: D.C.B., A.K.; investigation: D.C.B.; data curation: D.C.B., A.K., P.D.K, M.J.D., S.K., M.K.J.; writing: D.C.B., H.E., S.G., Z.K., M.K.J.; supervision: D.L.C., M.T.P., M.A.A., N.P.; reviewing & editing: all authors.

## Code Availability

All code used for processing data, figure generation, and statistics can be found in the publicly available GitHub repository: https://github.com/HOMES-Clinic/volunteer_outcomes_study.git

## Data Availability

Anonymized data used for this study are available from the corresponding author upon reasonable request.

