## Supplemental Figures 1-2 for "Volunteering at a Student-Run Clinic and Matching into Primary Care Specialties"

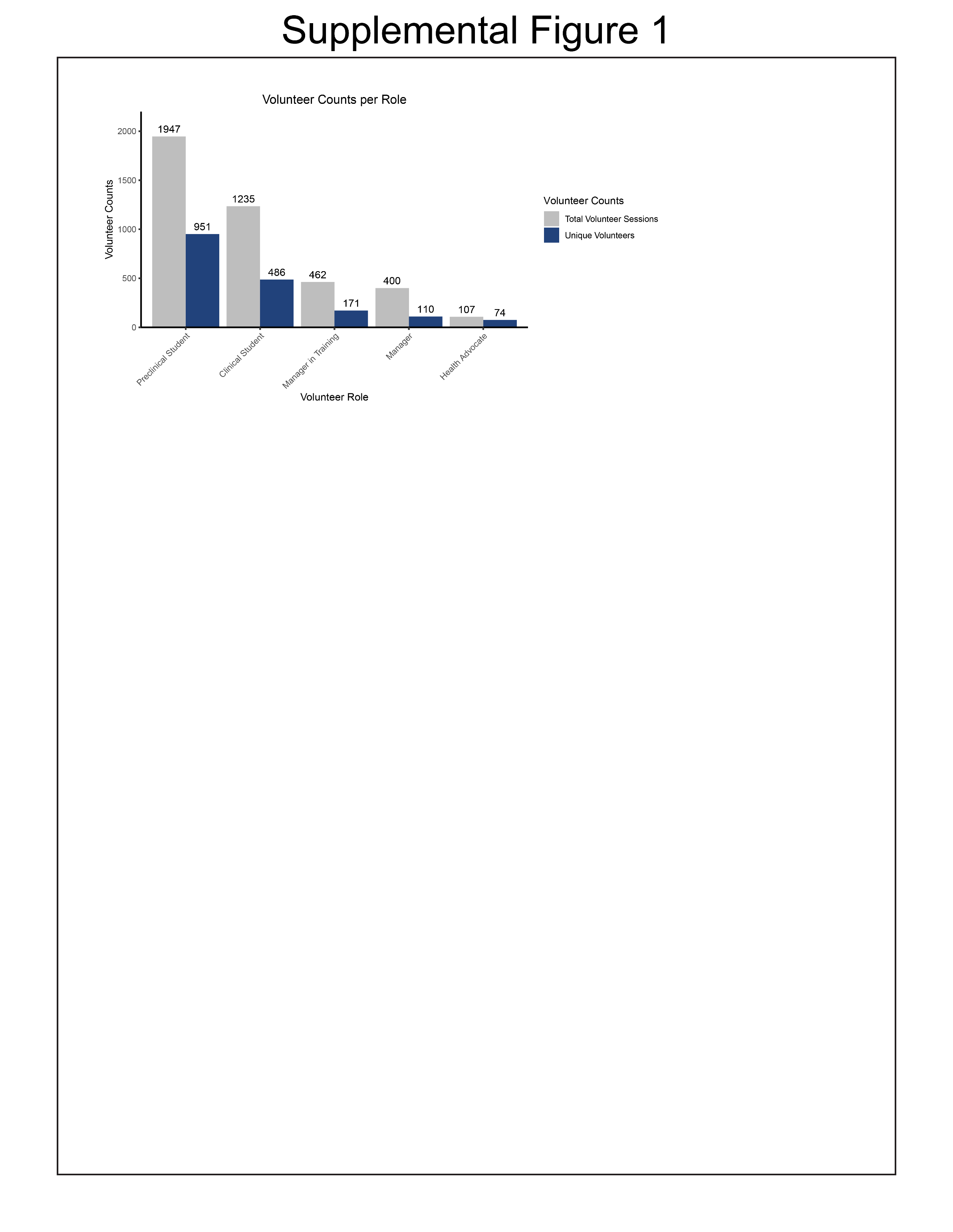

**Supplemental Figure 1: Volunteer Counts per Role.** Counts for the number of volunteer sessions and number of unique volunteers per clinic role.

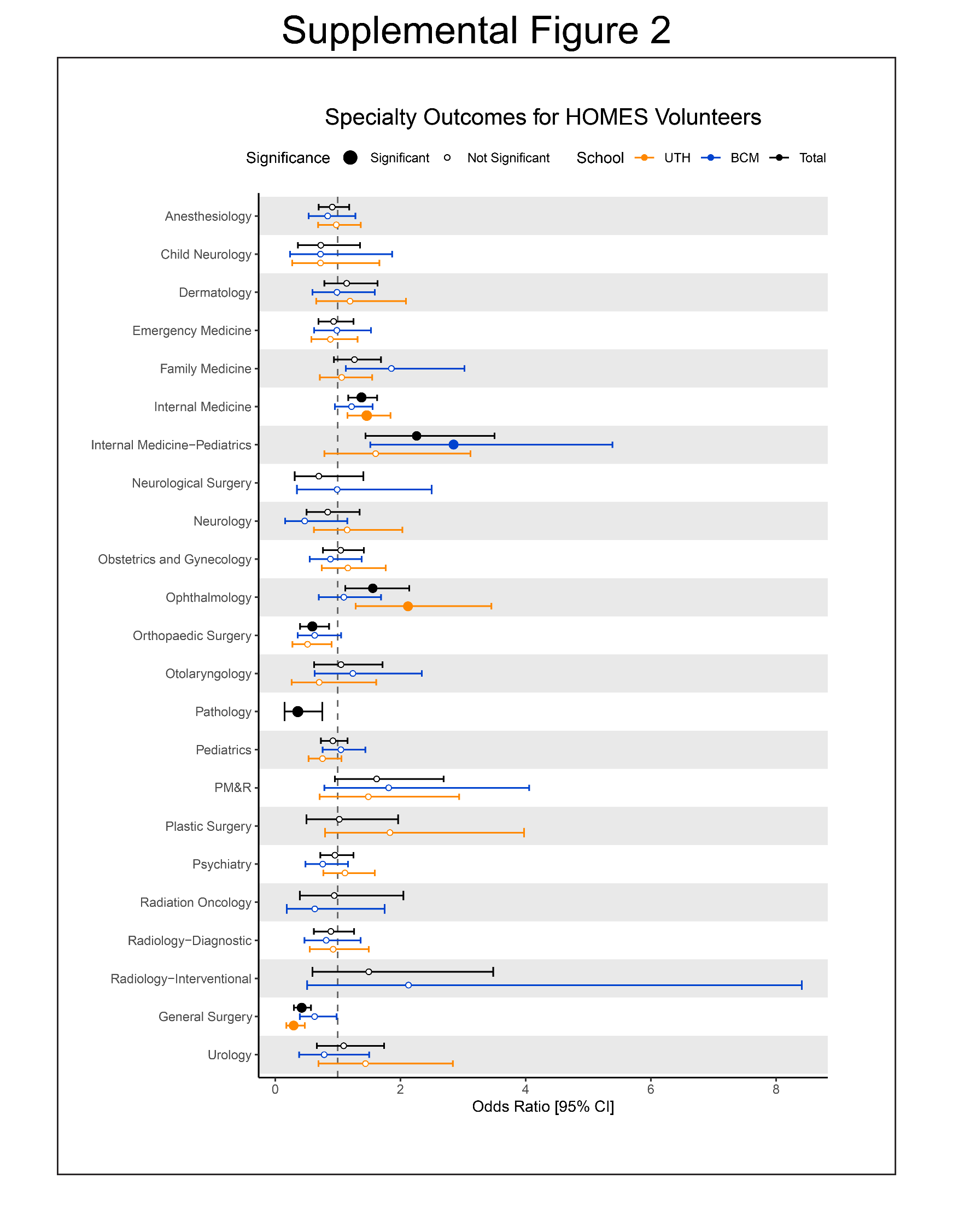

**Supplemental Figure 2: Match Outcomes of HOMES Volunteers Compared to Non-Volunteers by Affiliate Medical School.** Specialty-specific odds ratios are shown as points, where the error bars represent the 95% confidence interval. Statistical significance using FDR-adjusted p-values are denoted by a large solid point, while non-significant associations are denoted by hallow points. Affiliate medical school is represented by color, with BCM shown in blue, UTH shown in orange, and the composite association shown in black.
